# Predicting the Risk of Progression from Barrett’s Esophagus to Esophageal Adenocarcinoma with a Mass-Spectrometry-Based Proteomic Panel

**DOI:** 10.1101/2024.10.16.24315346

**Authors:** Andrew Cannon, Mark Hiatt, Igor Ban, Saed Sayad, Ted Karkus, Christopher Hartley

## Abstract

**Background:** Esophageal adenocarcinoma (EAC) is a highly aggressive cancer with poor prognosis, often arising from gastroesophageal reflux disease (GERD) and its precursor, Barrett’s esophagus (BE). Although only a small proportion of individuals with GERD or BE develop EAC, accurately identifying those at high risk is critical for early intervention. Current surveillance methods, such as endoscopy, are invasive and have limited accuracy in predicting malignant transformation. Mass-spectrometry-based proteomics offers a promising alternative by identifying protein biomarkers associated with disease progression.

**Method:** This study evaluated an eight-protein panel in predicting the risk of progression from BE to BE with high-grade dysplasia or EAC, potentially improving early detection and patient outcomes. A cohort of 107 subjects diagnosed with BE was recruited, and biological samples (tissue biopsies) were collected and labeled by a pathologist. Proteomic analysis was conducted using mass spectrometry to profile protein expression across the samples. A statistical test (t-test) was applied to assess differences in the proteomic panel across BE that progressed on follow-up compared to BE without progression.

**Results:** The t-test results revealed significant differences in protein expression levels between subjects with BE who progressed to cancer and those who did not. Key proteins, including CNDP2_TVF, DAD1 (detected via the FLE and ADF peptides), and GPI_LQQ, demonstrated strong statistical significance (p < 0.001) for under-expression, suggesting their potential as early biomarkers for identifying individuals at high risk. S100p_YSG also showed meaningful under-expression (p = 0.0111), indicating its relevance in cancer progression pathways. In contrast, LTF_DGA was over-expressed (p = 0.0226), possibly serving as an activator in the progression pathway and offering a novel therapeutic target.

**Conclusions:** This study identified promising biomarkers on the panel—including CNDP2_TVF, DAD1 (detected via the FLE and ADF peptides), and GPI_LQQ—associated with the development of advanced cancer in BE. Their under-expression could inform novel diagnostic and therapeutic approaches. Integrating these protein profiles with genetic and epigenetic data may provide a more complete understanding of disease progression. Longitudinal studies tracking protein levels over time may also reveal their potential as early cancer markers.

## Introduction

Esophageal adenocarcinoma (EAC) is a highly aggressive malignancy with a dismal prognosis, often due to late-stage diagnosis. Gastroesophageal reflux disease (GERD) and Barrett’s esophagus (BE) are well-established precursors, with chronic reflux leading to metaplasia of normal esophageal tissue into intestinal-type epithelium (Figure 1). While a small subset of patients with GERD or BE progress to EAC, accurately identifying those at high risk is paramount for timely intervention and improved outcomes. Traditional surveillance methods, such as endoscopy and biopsy, have limitations in predicting progression to EAC. Biomarkers are urgently needed to stratify patients based on their risk of developing this malignancy. Mass-spectrometry-based proteomics offers a promising avenue, enabling the identification and quantification of a multitude of proteins associated with disease progression.

**Figure 1:**
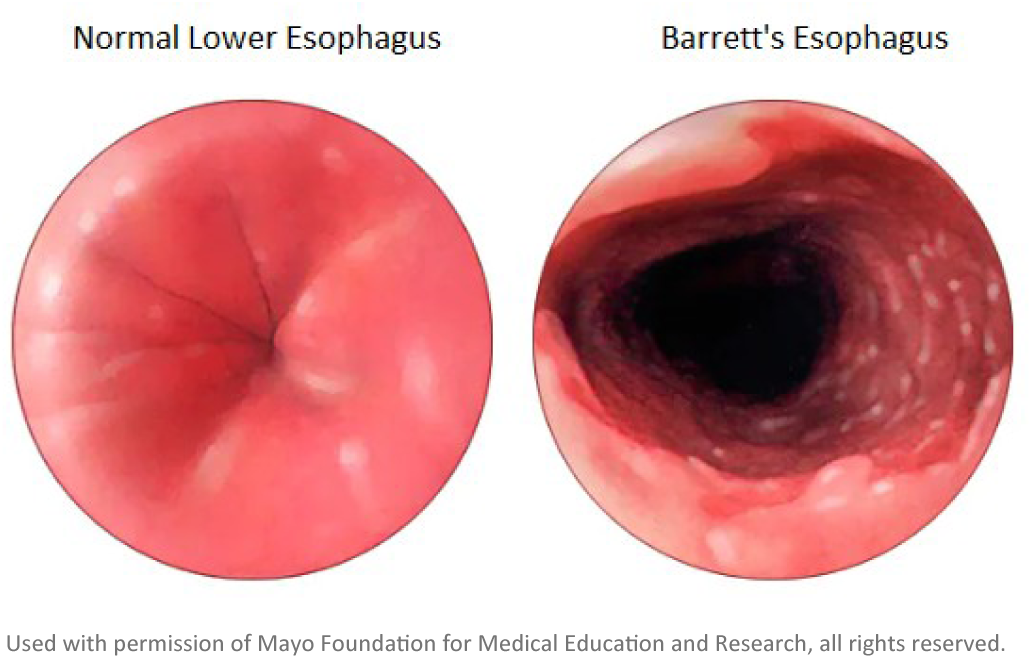
Normal and Barrett’s esophagus.

This study evaluated an eight-protein panel (BE-Smart, ProPhase Labs, Garden City, NY)^1^ in predicting
the risk of progression from BE to EAC. By identifying key biomarkers through mass spectrometry and
their association with disease progression, we assessed the feasibility of a means for early risk stratification. Such a tool could reduce the incidence and mortality of EAC through earlier diagnosis and targeted monitoring. Also, the majority of patients who do not develop EAC following a diagnosis of BE could be spared numerous endoscopies, reducing procedure morbidity and healthcare costs.

## Data

One hundred seven formalin-fixed paraffin-embedded tissue samples from patients diagnosed with BE were provided by Mayo Clinic (Rochester, MN), as approved by its institutional review board. These samples were processed using Liquid Tissue™ (Expression Pathology Inc., Rockville, MD) to allow for proteomic analysis. To profile protein expression, the samples were then analyzed using a highresolution mass spectrometer by mProbe, Inc. (Rockville, MD), whose laboratory was accredited by the College of American Pathologists and certified under the Clinical Laboratory Improvement Amendments. Among the 107, 73 patients did not progress to high-grade dysplasia or adenocarcinoma, while 34 patients did advance to these conditions (Figure 2). This cohort may be divided into three categories:(1) Barrett’s Patients with No Dysplasia in the Esophagus, (2) Barrett’s Patients with Low Dysplasia in the Esophagus, and (3) Barrett’s Patients with Indefinite Dysplasia in the Esophagus (Figure 3).

**Figure 2:**
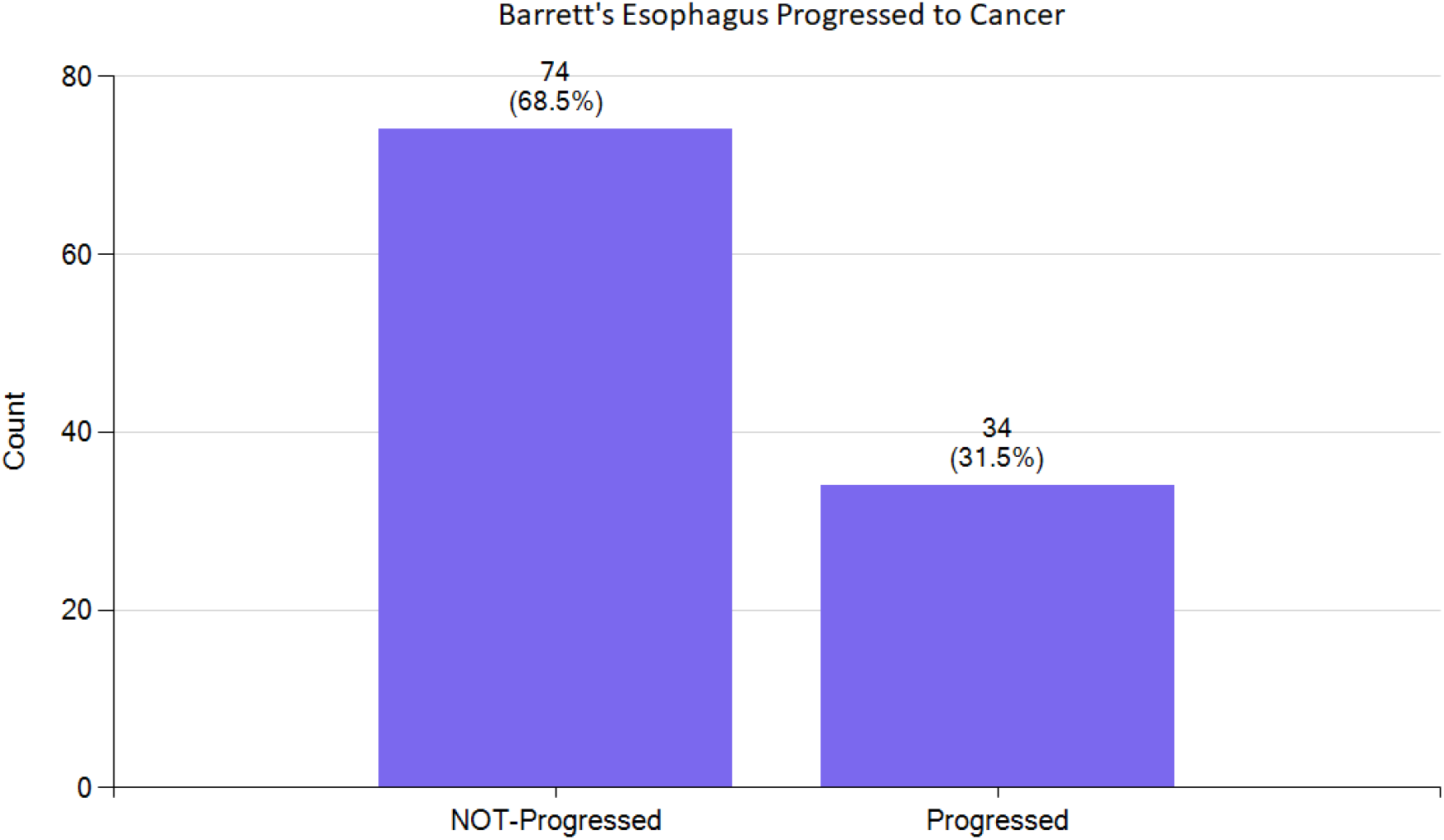
Of 107 patients, 74 did not progress to high-grade dysplasia or adenocarcinoma, while 34 did progress to these conditions.

**Figure 3:**
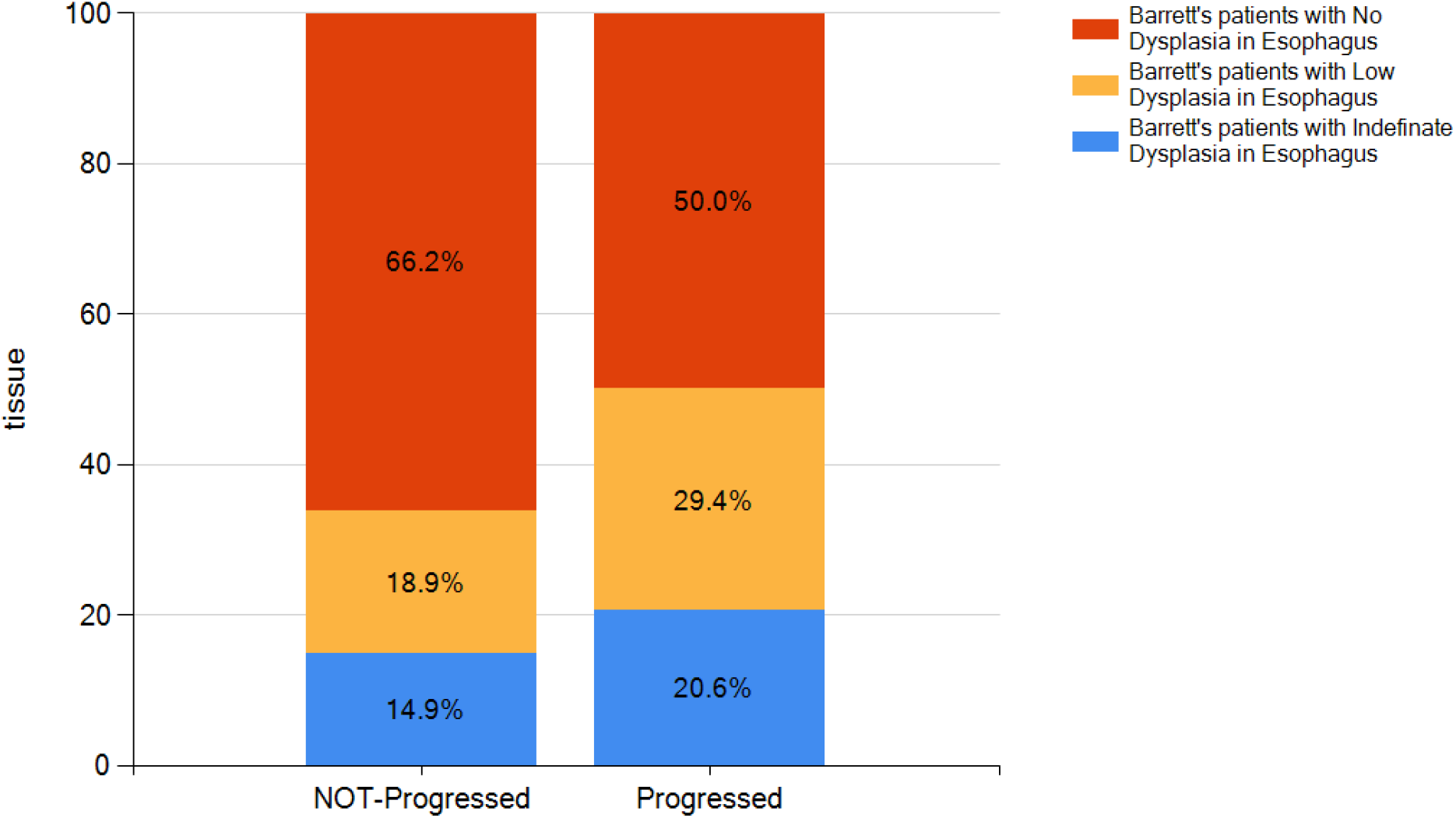
The cohort comprised of 107 patients diagnosed with BE may be divided into three categories.

## Statistical Analysis

Proteomic analysis was conducted using mass spectrometry to profile protein expression across the samples. Using the Genomarker platform (Bioada Lab, Toronto, ON), a statistical test (t-test) was applied to assess differences in the proteomic panel across BE that progressed on follow-up compared to BE without progression. Table 1 summarizes these findings, highlighting proteins that were under- or over-expressed in progressed patients, with lower p-values indicating stronger statistical significance.

**Table 1:** Statistical analysis of protein expression in subjects with BE. *(In naming these proteins, the string before the underscore* denotes *the parent protein and the string following the underscore denotes the proteotypic peptide used to quantify the parent protein.)*

| Protein | P-value (t-test) | Expression in Progressors |
| --- | --- | --- |
| CNDP2_TVFL | 0.00002 | Under-expressed |
| DAD1_FLE | 0.0003 | Under-expressed |
| GPI_LQQ | 0.0003 | Under-expressed |
| DAD1_ADF | 0.0004 | Under-expressed |
| S100p_YSG | 0.0111 | Under-expressed |
| CNDP2_LP | 0.0195 | Under-expressed |
| LTF_DGA | 0.0226 | Over-expressed |
| IGS15_IGV | 0.0655 | Under-expressed |
| UBE2N_YFH | 0.0904 | Under-expressed |
| S100p_ELP | 0.1019 | Under-expressed |
| LTF_FQL | 0.2219 | Over-expressed |
| IGS15_LAV | 0.2750 | Under-expressed |
| SET_LNE | 0.6113 | Under-expressed |

### CNDP2_TVF and CNDP2_LP

Carnosine Dipeptidase 2 (CNDP2) is an enzyme involved in breaking down dipeptides, such as carnosine, that protect cells from oxidative stress. CNDP2 acts as a functional tumor suppressor in gastric cancer through activating the mitogen-activated protein kinase (MAPK) pathway [1]. Its under-expression may result in less oxidative protection and increased susceptibility to DNA damage, potentially contributing to cancer progression in high-risk tissues such as those in BE [2].

### DAD1_FLE and DAD1_ADF

Defender Against Cell Death 1 (DAD1), an anti-apoptotic protein essential for cell survival, is associated with cancer initiation and progression. It has been demonstrated that DAD1 plays critical roles, including as a diagnostic or prognostic biomarker, in several types of cancer and performs various key functions, such as participating in cellular apoptosis, invasion, and chemosensitivity [3].

### GPI_LQQ

Glucose-6-Phosphate Isomerase (GPI) is a multifunctional enzyme that catalyzes the conversion of glucose-6-phosphate to fructose-6-phosphate in glycolysis. GPI supports cellular metabolism, especially in cancer cells that rely heavily on GPI glycolysis for energy (as in the “Warburg effect”) [4]. GPI has been linked to the proliferation and motility of cancer cells through its control over glucose-6-phosphate levels.

### S100p_YSG and S100p_ELP

S100P, a member of the S100 family of small calcium-binding proteins, can function both inside and outside cells. It has been found to be over-expressed in various cancers, in which it is linked to drug resistance, metastasis, and poor patient outcomes [5]. S100P under-expression could influence inflammatory and proliferative pathways, possibly reducing its effects on cellular signaling pathways that cancer cells often exploit for rapid growth and metastasis.

### LTF_DGA and LTF_FQL

Lactotransferrin (LTF) is an iron-binding glycoprotein with roles in immune function, including antimicrobial, anti-inflammatory, and anti-tumor activities. It can help regulate the availability of iron, which cancer cells need for growth. Over-expression of LTF may enhance iron regulation and support an immune environment less favorable to cancer. However, excess iron regulation may also be favorable to cell proliferation in certain cancer contexts, indicating that LTF’s role in oncogenesis could vary depending on the tissue and type of cancer. These findings suggest that the LTF protein family may play a crucial role in the progression from BE to EAC [6].

### IGS15_IGV and IGS15_LAV

Interferon-stimulated genes (ISGs), induced by type I interferons, play a crucial role in regulating immune responses against pathogens. Among these, ISG15 is especially noteworthy for its dual role in cancer, in which it can act as either pro- or anti-tumorigenic depending on the context. ISG15 also modulates the tumor microenvironment by influencing immune cell infiltration, inflammation, and tumor progression or suppression. Its dysregulation in various cancers highlights its diverse impact across cancer types, presenting ISG15 as a promising therapeutic target for enhancing anti-cancer immune responses or modulating tumor growth in cancer treatments [7].

### UBE2N_YFH

Ubiquitin-conjugating enzyme E2 N (UBE2N) is part of the ubiquitin-proteasome pathway, which regulates protein degradation. The ubiquitin-mediated degradation system plays a key role in regulating tumor-promoting processes such as DNA repair, cell cycle arrest, cell proliferation, apoptosis, angiogenesis, migration, invasion, metastasis, and drug resistance. Ubiquitin is attached to target proteins through a sequential cascade involving E1 (activating), E2 (conjugating), and E3 (ligating) enzymes, with E2 enzymes serving as central regulators within the ubiquitination system, influencing multiple pathophysiological processes in the tumor microenvironment [8].

### SET_LNE

Su(var)3-9, Enhancer-of-zeste and Trithorax are all proteins that contain a SET domain, which is associated with histone methyltransferase activity, allowing them to modify chromatin and regulate gene expression. Their role in cancer involves epigenetic regulation, in which they contribute to the activation or silencing of genes involved in cell growth, differentiation, and apoptosis [9].

## Discussion

The results of the statistical analysis between subjects with BE who progressed to cancer and those who did not revealed significant differences in the expression levels of several proteins. In particular, proteins CNDP2_TVF, DAD1 (detected via the FLE and ADF peptides), and GPI_LQQ demonstrated strong statistical significance, with p-values below 0.001, indicating that they were significantly under-expressed in patients who progressed to cancer. These findings suggest a role for these proteins in the
early identification of patients at higher risk for progression, given the clear differential expression pattern observed.

Furthermore, S100p_YSG showed a statistically significant under-expression (p = 0.0111), positioning it just above the highly stringent threshold of 0.01, but still within the range considered clinically meaningful. This under-expression, along with the other highly significant proteins, suggests that suppression of these proteins may be linked to pathways involved in cancer progression within BE. The mechanism underlying their under-expression warrants further investigation to determine if these proteins could be therapeutic targets to halt progression.

The protein LTF_DGA stands out as it is over-expressed in subjects who progressed to cancer, with a moderately significant p-value of 0.0226. This over-expression suggests a different role for LTF_DGA compared to the primarily under-expressed proteins, potentially acting as an activator or facilitator in the progression pathway. If further studies confirm this trend, LTF_DGA could serve as an additional biomarker and a novel target for therapeutic intervention aimed at slowing or preventing cancer progression.

Proteins with non-significant p-values—such as IGS15_IGV, UBE2N_YFH, and SET_LNE—did not exhibit a statistically meaningful difference between the two subject groups, indicating that they may not be directly involved in the progression of BE to cancer. This finding suggests that not all proteins in the study contribute to the disease’s progression, which is essential in narrowing down a focused list of biomarkers.

## Summary

This study identified key proteins with differential expression in patients progressing to cancer, shedding light on the biological mechanisms underlying cancer development in BE. Notable biomarkers include CNDP2_TVF, DAD1 (detected via the FLE and ADF peptides), and GPI_LQQ, whose under-expression is linked to advanced cancer stages in BE and may guide the development of novel diagnostic and therapeutic strategies. In particular, the BE-Smart proteomic panel is comprised of predictive biomarkers that are both statistically significant and mechanistically meaningful. Combining these protein profiles with genetic and epigenetic data could enhance our understanding of disease progression. Additionally, longitudinal studies monitoring protein levels over time may further elucidate their potential as early indicators of cancer.

## Data Availability

All data produced in the present study are available upon reasonable request to the authors

## Footnotes

1 BE-Smart contains the following eight proteins: CNDP2, DAD1, GPI, ISG15, LTF, S100P, SET, and UGE2N.

## Notes

### Competing Interest Statement

The authors have declared no competing interest.

### Funding Statement

This study did not receive any funding

### Author Declarations

Mayo Clinic IRB IRB Application #: 23-000689 IRB Approval Date: 3/9/2023 Christopher Hartley Makala Amundson Andrew Cannon Catherine Hagen Christopher Hartley Priyadharshini Sivasubramaniam

### Summary of Updates

We have included the permission line for Figure 1 in the manuscript following receipt of the permission letter from the Mayo Clinic.

## References

1. Zhang Z, Miao L, Xin X, Zhang J, Yang S, Miao M, Kong X, Jiao B. Underexpressed CNDP2 participates in gastric cancer growth inhibition through activating the MAPK signaling pathway. Mol Med. 2014 Mar 13;20(1):17–28.

2. Prokopieva VD, Yarygina EG, Bokhan NA, Ivanova SA. Use of Carnosine for Oxidative Stress Reduction in Different Pathologies. Oxid Med Cell Longev. 2016;2016:2939087.

3. Luo Y, Wu Y, Huang H, Yi N, Chen Y. Emerging role of BAD and DAD1 as potential targets and biomarkers in cancer. Oncol Lett. 2021 Dec;22(6):811.

4. Liberti MV, Locasale JW. The Warburg Effect: How Does it Benefit Cancer Cells? Trends Biochem Sci. 2016 Mar;41(3):211–218. doi: 10.1016/j.tibs.2015.12.001. Epub 2016 Jan 5. Erratum in: Trends Biochem Sci. 2016 Mar;41(3):287. Erratum in: Trends Biochem Sci. 2016 Mar;41(3):287.

5. Arumugam T, Logsdon CD. S100P: a novel therapeutic target for cancer. Amino Acids. 2011 Oct;41(4):893–9.

6. Rascón-Cruz Q, Espinoza-Sánchez EA, Siqueiros-Cendón TS, Nakamura-Bencomo SI, Arévalo-Gallegos S, Iglesias-Figueroa BF. Lactoferrin: A Glycoprotein Involved in Immunomodulation, Anticancer, and Antimicrobial Processes. Molecules. 2021 Jan 3;26(1):205.

7. Nguyen HM, Gaikwad S, Oladejo M, Agrawal MY, Srivastava SK, Wood LM. Interferon stimulated gene 15 (ISG15) in cancer: An update. Cancer Lett. 2023 Mar 1;556:216080.

8. Bui QT, Hong JH, Kwak M, Lee JY, Lee PC. Ubiquitin-Conjugating Enzymes in Cancer. Cells. 2021 Jun 4;10(6):1383.

9. Yang Z, Zhang C, Che N, Feng Y, Li C, Xuan Y. Su(var)3-9, Enhancer of Zeste, and Trithorax Domain-Containing 5 Facilitates Tumor Growth and Pulmonary Metastasis through Up-Regulation of AKT1 Signaling in Breast Cancer. Am J Pathol. 2021 Jan;191(1):180–193.

